# CLASS - CiaoLApo Stillbirth Support checklist: adherence to stillbirth guidelines and women’s psychological well-being

**DOI:** 10.1101/2023.06.07.23291084

**Authors:** Claudia Ravaldi, Francesca Roper, Laura Mosconi, Alfredo Vannacci

## Abstract

**Background:** Stillbirth is a global public health issue affecting millions of parents and healthcare professionals. Quality and consistency of bereavement care after stillbirth are crucial for parents’ well-being, but they depend on the implementation and impact of international guidelines.

**Aim:** This study aimed to validate practices for stillbirth care by means of the CiaoLapo Stillbirth Support (CLASS) checklist, a tool that summarises the common elements of the main international guidelines on bereavement care after stillbirth, and to explore their association with women’s satisfaction, respectful care, and psychological outcomes.

**Methods:** A cross-sectional online survey was conducted among 261 women who experienced a stillbirth in the last 10 years in Italy, a country without official national guidelines. The survey included the CLASS checklist, the Perinatal Grief Scale, the National Stressful Events Survey PTSD Short Scale, and questions on satisfaction and respectful care.

**Findings:** The mean score of adherence to guidelines was low, 2.0 (SD 1.1, on a 4-point scale), with regional differences. The lowest scores were in respect for the baby, communication about funeral and autopsy, creating memories, and aftercare. Adherence to at least 40 out of 60 CLASS checklist items was independently associated with greater satisfaction and respectful care, and lower grief and posttraumatic stress symptoms.

**Conclusion:** The study shows that women who receive care that adheres to stillbirth guidelines have a better psychological outcome, with lower levels of grief and posttraumatic stress symptoms. The study also highlights the need for official national guidelines and adequate training and support for healthcare professionals in countries where they are lacking.

## 1 Introduction

Each year, 2.6 million pregnancies worldwide end in stillbirths, most of which are preventable (1). In the last decade, this number has mostly remained unchanged (1). This is a demonstration of the fact that stillbirths are still deeply deprioritized globally and nationally (2). Stillbirth is mostly invisible as a global health issue, and on a national level many countries do not have public health plans to reduce preventable stillbirths and do not collect or produce quality stillbirth data (3). Aside from impacting mortality rates and affecting women’s physical health, stillbirths can heavily impair women’s mental health (2,4). Suffering a stillbirth is a traumatic event for women and their partners (5). If not cared for in a supportive and respectful manner, a stillbirth can lead to adverse psychological outcomes, which can ultimately lead to an increased burden on society (6). However, equitable access to high-quality bereavement care following a stillbirth is still a neglected component of healthcare services (2). Additionally, providing bereavement care can have negative psychological impacts on healthcare professionals, such as professional burnout, if they are untrained and emotionally unprepared and left alone to deal with such stressful events (7).

Given the impact of stillbirths on both parents and healthcare professionals, guidelines aimed at improving the quality of assistance following a stillbirth are necessary and extremely important (7,8). The first studies on the clinical management of stillbirths, aiming to facilitate normal mourning processes, date back to the 1970s (9). Such studies were written in a context where healthcare professionals, due to the silence and stigma surrounding the topic, were reluctant to face the problem of stillbirth. Since then, also thanks to the contribution of the American and British bereaved parents associations, many countries have adopted national guidelines to improve assistance to women and partners affected by stillbirth. The main countries that have worked on national stillbirth guidelines are Australia and New Zealand (PSANZ - The Perinatal Society of Australia and New Zealand), Canada (Canadian Pediatric Society), the United Kingdom (NBCP - The National Bereavement Care Pathway, lead by SANDS -Stillbirth and Neonatal Death Charity), and Ireland (Health Services Executive Ireland) (10–13). The most relevant elements that are shared between these guidelines have been inserted in the CiaoLapo Stillbirth Support (CLASS) checklist, which has divided recommendations from the different guidelines into six sections: respect, information and communication, birth options, hospital stay, creating memories, and aftercare (8,14).

In many position statements WHO has called for the need of respectful care and official national guidelines to aid healthcare professionals in providing such respectful and supportive bereavement care (4,15). Yet, many countries around the world still do not have official national guidelines. Italy is one such country, where healthcare professionals are left without any formal guidelines when assisting stillbirths. Recently, Italian recommendations about clinical management of stillbirth have been published (16), through the joint work of some of the major Italian obstetricians’ associations. Suggestions about care are similar to those provided by international guidelines and summarised in the CLASS checklist. Although such recommendations are a first important step towards formal guidelines, they are far from being implemented in clinical practice as formal guidelines endorsed by the Italian National Health Service.

CiaoLapo Foundation, an Italian charity founded in 2006, has since its inception provided more than 800 hours of formal training to healthcare professionals in 100 maternity units (i.e. 20% of all Italian maternity units) (17). Additionally, many units have adopted the guidelines on bereavement care proposed by CiaoLapo, and have reported an improvement in the assistance to stillbirths, with parents assisted after the healthcare professionals’ training reporting a greater satisfaction of care when compared to bereaved parents assisted prior to the training (8,18,19). Despite such improvements, Italy is still facing two major issues: 1) most units still do not have local bereavement care procedures, and 2) the Italian national health service has not yet developed official national guidelines.

This situation, characterised by an uneven distribution of guidelines throughout the country, with varying standards of care over time even within the same facility, prompted us to investigate the effect of guidelines on bereavement assistance and on the psychological parameters of parents. Using the CLASS checklist, this study aims to explore how bereaved parents’ psychological wellbeing was influenced by respectful care received and compliance to international guidelines.

## 2 Methods

This study is part of the permanent OPALE project (Observatory on PerinatAL hEalth) and follows a cross-sectional study design. The survey is hosted on the Qualtrics platform provided by Florence University PeaRL laboratory, and is distributed through the online channels of CiaoLapo Foundation, an organisation working on the promotion of perinatal health in Italy. Information about the questionnaire was provided in the first page of the survey, and consent was given by participants before proceeding with the survey. Participants were selected for the CLASS study if they were female and had suffered a pregnancy loss after 20 gestational weeks (including termination of pregnancy due to pathology) in the last 10 years.

The following sections of the OPALE survey were used for the CLASS study:

1. Socio-demographic information and medical history.
2. The CLASS checklist, with 60 items common to the main international guidelines on bereavement care (10–13), covering the following six areas:
  a. <u>Respect for the baby and parents</u>, i.e., naming the baby, bathing and dressing the baby, providing privacy, allowing partners to spend time together, etc.
  b. <u>Information and communication</u>, i.e., using parent-friendly language, avoiding dehumanising terms and medical jargon, giving written information, discussing issues at most appropriate time in a quiet and private environment, etc.
  c. <u>Birth Options</u>, i.e., offering parents choices, offering the option of returning home, offering obstetric analgesia, avoiding sedation, etc.
  d. <u>Hospital stay</u>, i.e., providing privacy, allowing time with the baby, not urging parents to leave the hospital, etc.
  e. <u>Creating Memories</u>, i.e., allowing parents to see and hold their baby, providing mementos such as a lock of hair, footprints, ID bracelet, etc.
  f. <u>Aftercare</u>, i.e., informing mothers about physical and psychological consequences of perinatal loss, providing early psychological support, providing written information on support services, discussing implications for future pregnancies, etc.
  The CLASS checklist was used as a measure of adherence to international stillbirth guidelines. Parents were asked to rate every item from 0 (not respected at all) to 4 (fully respected) or to select the option “I don’t know or not applicable”. Items were considered “satisfied” with a score equal or greater than 2. The CLASS checklist is available online in Italian and English (www.class.ciaolapo.it); English version is provided here as **Supplementary Table 1**.
3. The Perinatal Grief Scale (PGS), an instrument used to assess the grief after perinatal loss (22,23). The scale consisted of 33 Likert-type questions, with answers ranging from 1 (strongly agree) to 5 (strongly disagree). The PGS presents three subscales: ‘active grief’ (AG), ‘difficulty in coping’ (DC) and ‘despair’ (D).
4. The National Stressful Events Survey PTSD Short Scale (NSESSS), a 9-item measure that assesses the severity of posttraumatic stress disorder in individuals aged 18 and above following an extremely stressful event or experience (24). Each item asks the individual receiving care to rate the severity of his or her posttraumatic stress symptoms during the past 7 days. Each item is rated on a 5-point scale (0=Not at all; 1=A little bit; 2=Moderately; 3=Quite a bit, and 4=Extremely). The total score can range from 0 to 36 with higher scores indicating greater severity of posttraumatic stress symptoms.
5. The survey also included items assessing women’s satisfaction with care, and women’s perception of respectful care. Items were rated from 1 (not satisfied at all) to 5 (fully satisfied) and then classified as yes/no when needed for dichotomous outcomes analysis (25,26).

### 2.1 Statistical analysis and data presentation

Survey responses were extracted from the OPALE dataset hosted in Qualtrics platform and imported into Stata/BE 17.0 (StataCorp) for statistical analysis. Categorical data were reported as frequencies and percentages and compared using the chi-squared test, whereas continuous data were reported as mean values with standard deviations (SD) or as median [quartiles] and compared using t-test or Kruskall Wallis and Mann Witney test. All results were considered to be statistically significant at p < 0.05. Responders ’ location mapped by regional areas across Italy, scores, correlation graphs and box plots were plotted using Tableau Desktop 2023.1 (Tableau Software, LLC).

### 2.2 Ethic Statement

Ethics approval for the Observatory on PerinatAL hEalth OPALE was obtained by Florence University Ethics committee (n.175 prot 0261728, n.189 prot 0333881). The survey was voluntary and anonymous, no personal data were recorded, and in no way it is possible to identify the single respondents. Informed consent was obtained from all participants. The data were acquired in adherence with GDPR regulations (General Data Protection Regulation, European Union 2016/679).

## 3 Results

### 3.1 Sociodemographic information and medical history

The sample consisted of 261 women, with the majority (70.9%) being under 40 years old. Regarding education, almost 80% of the women had some level of tertiary education. The time since the perinatal loss was fairly evenly distributed, with 25.3% experiencing loss less than 12 months prior to answering the questionnaire. The gestational age at loss was equally distributed between 20-25 weeks (33.3%), 26-36 weeks (35.2%), and 37-42 weeks (31.4%); 14.0% of them reported a termination of pregnancy for medical reasons. Over half of the women (54.0%) had subsequent pregnancies, and 10.7% were currently pregnant. In terms of medical history, coagulopathy (11.5%) and obesity (14.6%) were the most common disorders reported, while anxiety was the most prevalent psychological history (23.4%). Finally, the majority of women (72.8%) experienced only one loss, while 17.2% experienced 2-3 losses, and 10% experienced more than three. Full details of the sample are reported in **Supplementary Table 2**.

### 3.2 Adherence with the CLASS checklist

The mean score of mothers’ perception of adherence to guidelines was quite low, 2.0 (SD 1.1; range 0.2 to 4.0; median value 1.9 [1.0 ; 3.0]). A significant difference between regions was observed, with Northern Italy scoring 2.32 [1.26 ; 3.38], Central Italy 2.00 [1.08 ; 2.92], and Southern Italy 1.48 [0.40 ; 2.56] (**Figure 1**).

**Figure 1.**
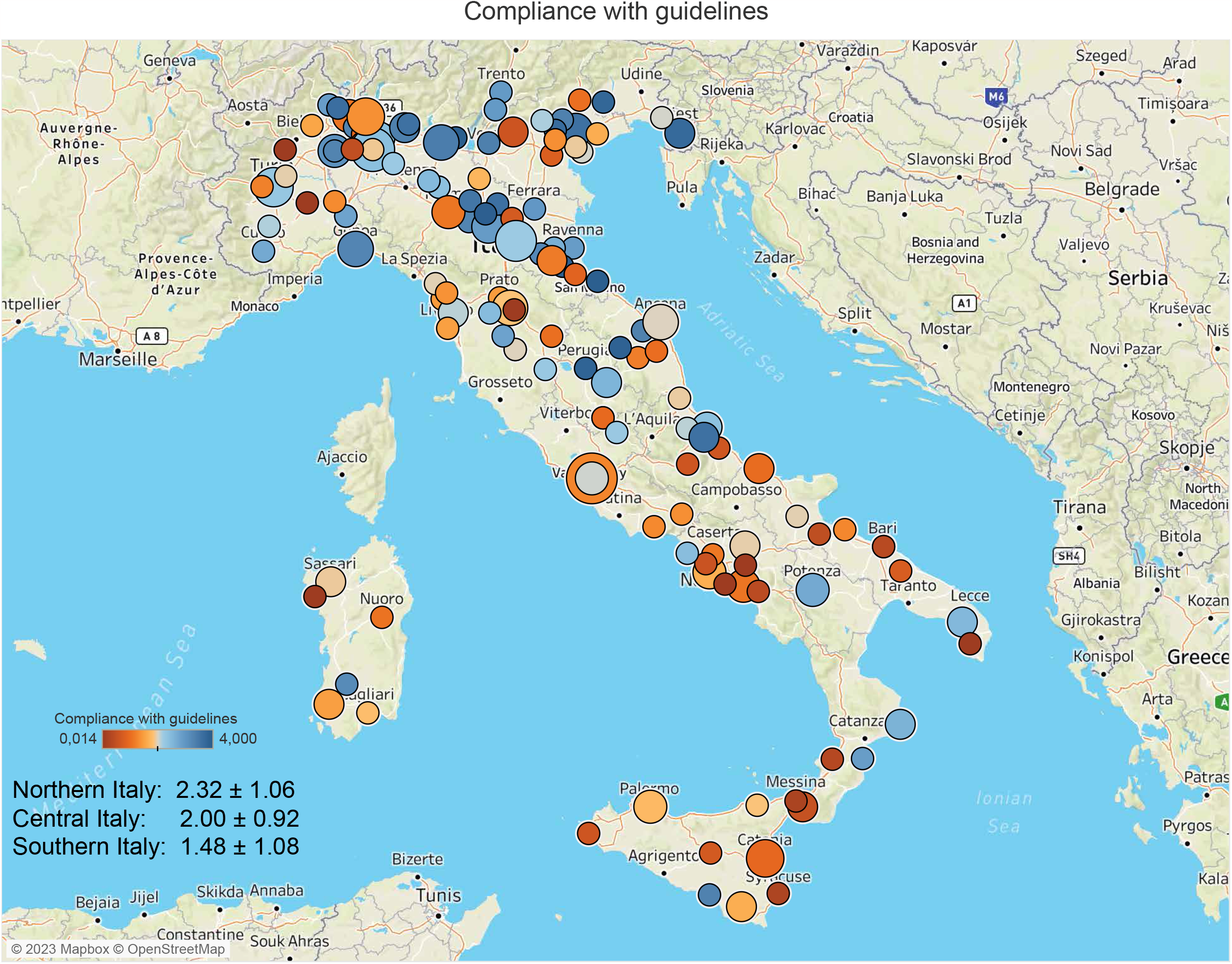

Mean scores of CLASS subcategories were also quite low (1.96 on a 4-point Likert scale), with the highest value (2.54, SD 1.2) in the “Mode of Birth” (Cb) section. Categories with ratings lower than the mean score were: A) respect, specifically towards the baby (Aa); B) communication, specifically about the baby’s funeral (Be) and autopsy (Bf); E) creating memories, particularly in relation to spending time with the baby (Ea), parenting (Eb), and having mementos (Ec); and F) aftercare, with regards to maternal changes (Fb), support services (Fc), and follow-up (Fd), with the lowest score reported in “Parenting” (Eb) section (0.61, SD 1.2) (**Figure 2**).

**Figure 2.**
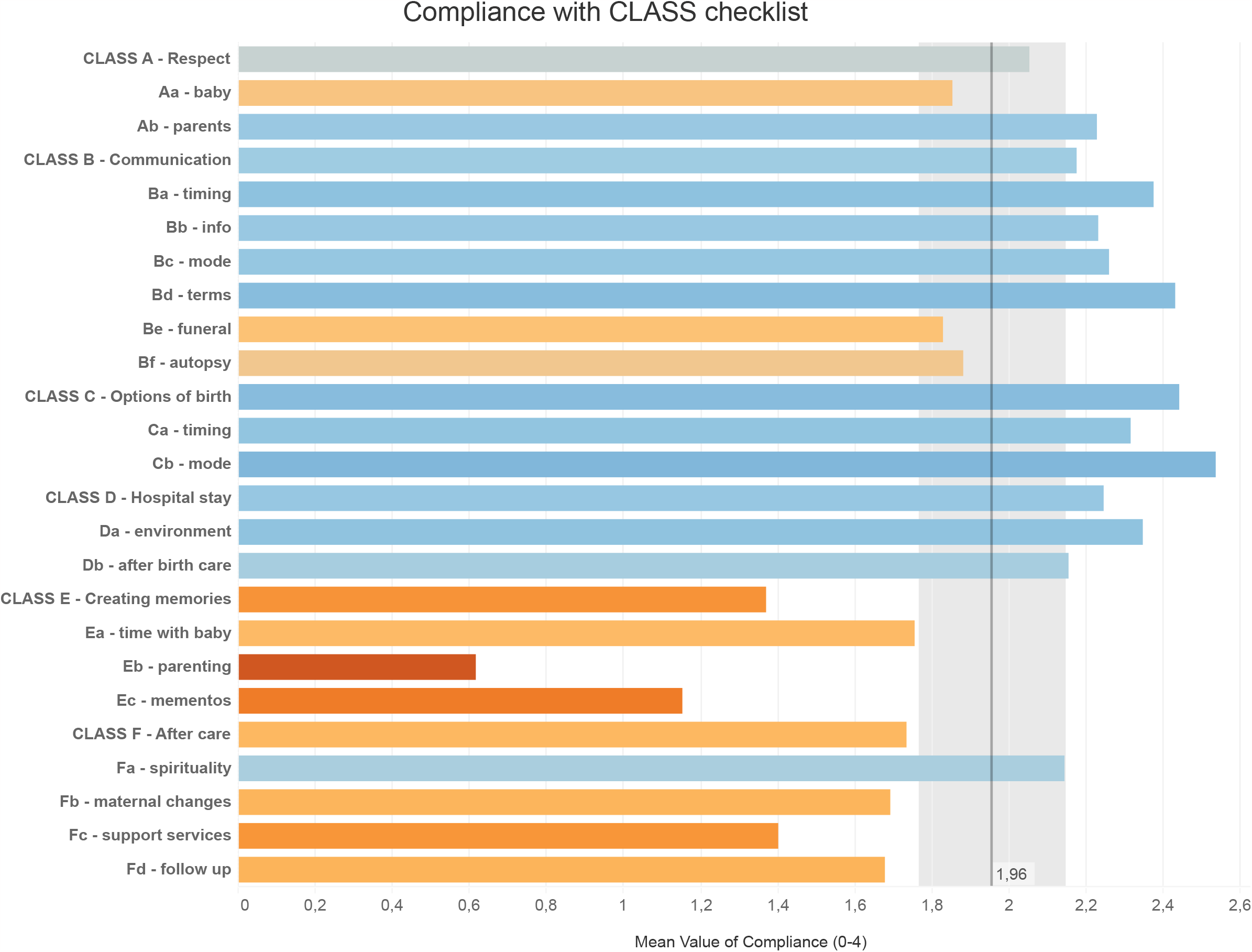

### 3.3 CLASS checklist, satisfaction, respectful care, and psychological outcomes

A higher mean score at the CLASS checklist was associated with a lower score at PGS and NSESSS (**Figure 3a**). Further, the more items of the CLASS checklist were satisfied, the less intensity of grief affected mothers (p<0.0001) (**Figure 3b**).

**Figure 3.**
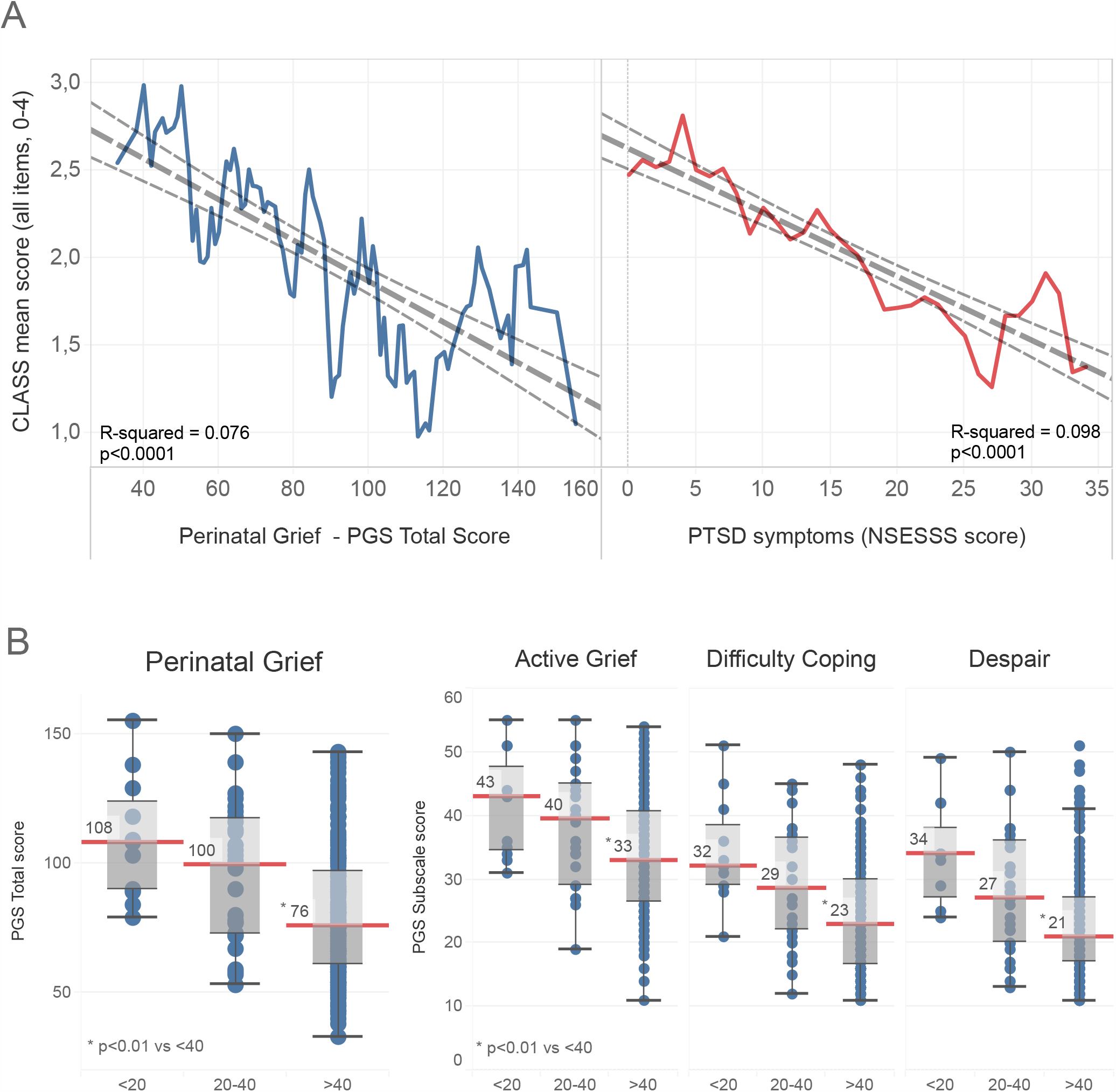

In a univariate analysis, all items of the CLASS checklist were found to be associated with mothers’ satisfaction of care (**Supplementary Figure 1**); in a multivariate analysis (**Figure 4**), adherence with at least 40 CLASS checklist items was found to be an independent factor related to greater satisfaction of care (OR 2.0, CI 1.1-3.8) and the perception of having received respectful care (OR 3.6, CI 1.9-7.0). A gestational age at loss between 20 and 25 weeks is positively associated with less satisfaction of care (OR 0.43, CI 0.2-0.9). Whereas, time after loss under 12 months is independently associated with higher satisfaction of care (OR 2.39, CI 1.1-4.9).

**Figure 4.**
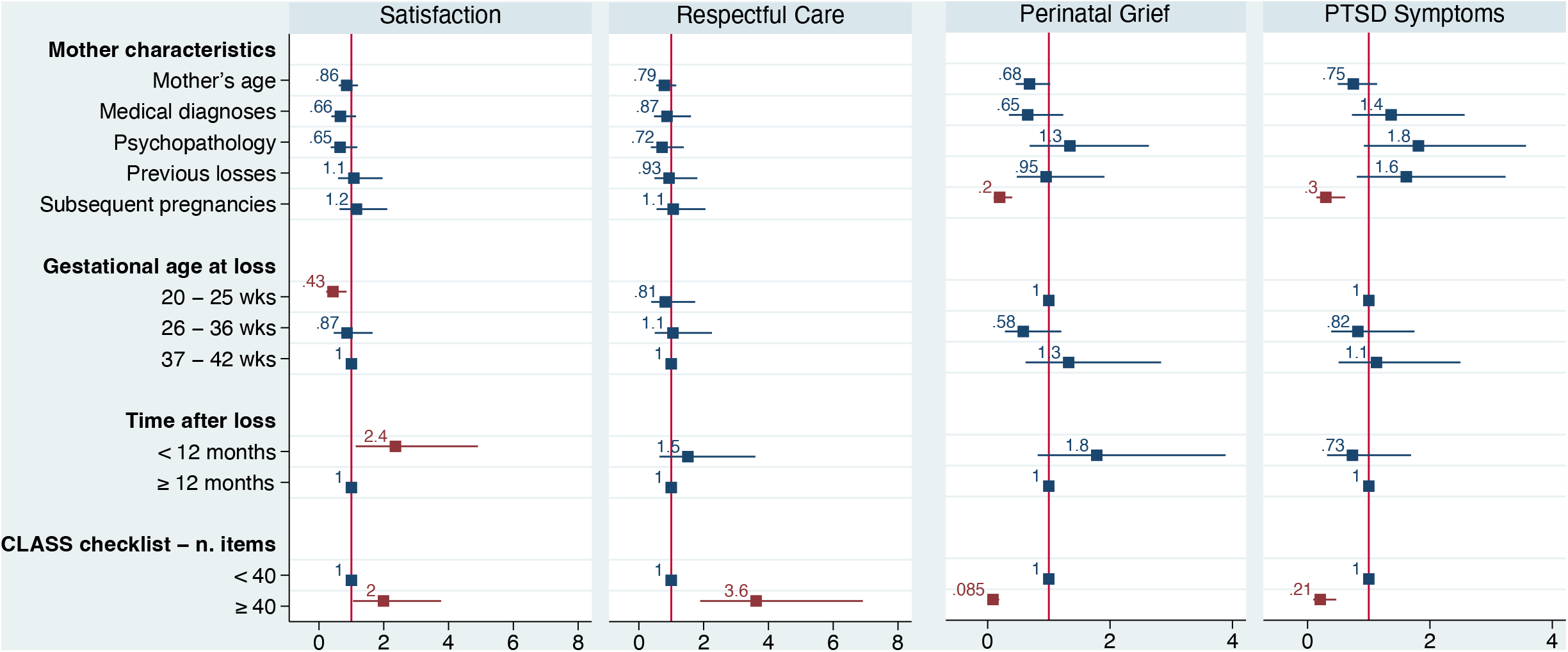

Adherence with at least 40 CLASS checklist items was also found to be an independent factor related to a lower score at PGS (OR 0.08, CI 0.1-0.2) and NSESSS (OR 0.21, CI 0.1-0.5). Also having a subsequent pregnancy after loss was related to a lower score at both scales (PGS -OR 0.20, CI 0.1-0.4; NSESSS -OR 0.30, CI 0.1-0.6).

## 4 Discussion

The primary objective of our research was to validate the internationally recognized guidelines on bereavement care following stillbirth, with a specific focus on bereaved parents’ perspectives and psychological well-being. To achieve this, we utilised the CLASS checklist, which encompasses recommendations from leading global guidelines, to evaluate parents’ understanding and opinions on the implementation of respectful care components. Furthermore, our secondary aim was to assess the current state of bereavement care in Italy. In order to accomplish these goals, we employed validated scales and open-ended questions to gauge parents’ satisfaction with care, their perception of respectful care, and their psychological outcomes. Our ultimate goal was to determine if adherence to these guidelines was correlated with improved outcomes for parents, thereby validating the effectiveness of the guidelines and providing insights into areas for improvement within the Italian context.

### 4.1 A snapshot of bereavement care

Our quantitative data showed that the mean score of mothers’ perception of adherence to guidelines was quite low (2.0 on a 4-point Likert scale), with significant regional differences. The most deficient categories were respect towards the baby, communication about funeral and autopsy, creating memories, and aftercare. Creating memories and respect towards the baby, which is closely linked to mementoes and memories itself, are acknowledged to be healing practices that facilitate the expression of grief and, consequently, the grieving process (27). However, this aspect, which is a crucial one, was often neglected or discouraged by healthcare professionals in Italy.

Aftercare services are also a crucial aspect of care pathways. Prior research has indicated that bereaved parents anticipate scheduled follow-up visits that offer not only medical consultation but also psychological support (28,29). It is possible that the lack of awareness among Italian healthcare professionals regarding these issues may be due to insufficient specialised training and the absence of relevant Italian guidelines on the subject (18,30).

### 4.2 Adherence to guidelines and psychological outcomes

The CLASS study effectively demonstrated the relationship between adherence to international guidelines, women’s psychological well-being, and satisfaction with care. Notably, adherence to 40 or more CLASS checklist items was associated with increased satisfaction and perception of respectful care, as well as reduced levels of grief and post-traumatic stress symptoms. This relationship is independent of potential confounding variables such as medical and psychological history, gestational age, elapsed time, and the presence of other children. Furthermore, a linear relationship was observed between adherence to guidelines and psychological outcomes, indicating that higher adherence corresponded to improved parental well-being.

Our findings also highlighted two distinct aspects: a lower satisfaction among women with a perinatal loss before 25 weeks of gestational age and a decrease in satisfaction as the time since the loss increases. We hypothesise that the first observation may be attributed to healthcare professionals potentially underestimating the psychological impact of an early loss on women. This hypothesis is supported by Rowlands and Lee (2009), in their paper “The silence was deafening”. Insensitive comments addressed to grieving mothers by healthcare professionals are described and they underpinned a lack of knowledge about the emotional aspects involved in early losses (32). In another paper, women perceived that staff didn’t understand what they were experiencing, again, underestimating women’s grief (33).

Regarding the second observation, as the duration since the loss increases, it is speculated that the initially experienced shock begins to diminish in the days or months following the event. During this period, women may seek information about perinatal grief, thereby gaining an understanding of the support they lacked during their hospital stay. This increased awareness may contribute to a decrease in overall satisfaction.

### 4.3 Guidelines and bereavement care

The low level of adherence to international guidelines reported by our sample reflects the lack of official national guidelines and the variability of care across different regions and units. This is consistent with previous studies that have documented the gaps and challenges in providing quality bereavement care in Italy (8).

Having national guidelines is the first step to guarantee respectful care, as stated by WHO (4,15). All healthcare professionals involved in bereavement care should accurately implement recommendations to assure the consistency of care. Inconsistency of bereavement interventions is still an issue to be faced which could undermine satisfaction of care and, consequently, the parents’ well-being (34). The recent publication of Italian recommendations on the clinical management of stillbirth (16), came after the data for this study was collected. This situation allowed us to compare parents who received low-quality care with those who received adequate care, all within similar social and cultural contexts and often even in the same hospital, since the quality of assistance often varied depending on the attending staff. In countries where national guidelines have been in place for years, a more homogeneous quality of care may exist, making comparisons between different levels of assistance more challenging.

Contemporary bereavement care requires healthcare professionals to possess basic knowledge in various disciplines, such as psychiatry, psychology, and sociology. To that end, a comprehensive and easy-to-use checklist based on the current literature can significantly aid in providing respectful and supportive assistance. The CLASS checklist, which incorporates the latest international recommendations, serves as a valuable tool in this context. Moreover, the CLASS study contributes to the validation of international guidelines on bereavement care by offering additional evidence of their crucial role for both parents and healthcare professionals.

### 4.4 Implications

This is the first study using clinical outcomes to show a significant and proportional relationship between adherence to stillbirth guidelines and psychological well-being of bereaved parents. The more items of the guidelines were met, the better the parents’ mental health, after adjusting for other potential confounding factors.

Therefore, we propose the use of the CLASS checklist as a tool to evaluate adherence with guidelines and promote perinatal mental health after perinatal loss. The CLASS checklist can help healthcare professionals to assess and improve their care quality, as well as to identify areas for further training and support. Moreover, the CLASS checklist can facilitate cross-cultural comparisons and collaborations among different countries and settings, as it incorporates the common elements of the main international guidelines on bereavement care after stillbirth. By using the CLASS checklist, we can advance our knowledge and understanding of the best practices and interventions to support parents and healthcare professionals after perinatal loss.

Finally, the CLASS study supports the need for official national guidelines on bereavement care after stillbirth in all countries that still lack them. Such guidelines would help standardise care across different regions and units, ensure quality and equity of care for all bereaved parents, and promote accountability and evaluation of care provision. Moreover, national guidelines would raise awareness and recognition of stillbirth as a public health issue that deserves attention.

## 5 Strengths and limitations

External review and validation of international guidelines represents the main strength of this study. The variety of bereavement care across Italy could at first glance be considered a limitation, but, from a merely methodological point of view, it should eventually be considered a strength. As a matter of fact, this inconsistency of care has allowed us to evaluate the impact of adherence to international guidelines on mothers’ well-being.

The wide distribution of the sample, in relation to time elapsed from loss, could entail a recall bias among mothers who had a stillbirth many years ago. However, it is known that in case of traumatic experiences the coherence of memories is quite well preserved (21), furthermore, this wide distribution allowed us to include more mothers which received different quality of care. Over the years, Italian bereavement care has been implemented, as shown in previous papers (7,8,18), and this enhancement has led to identify sample’s differences in terms of psychological outcomes. Through a multivariate analysis we have corrected the variable “time elapsed from loss” and, as shown in the results section, no difference was identified in mothers with a time elapsed from loss over 12 months in terms of psychological outcomes and satisfaction of care, suggesting that recall bias was minimal.

Another limit is that our study focused on high-income westernised countries as the source of international guidelines, which may not capture the cultural diversity and specific needs of parents from different backgrounds. Anyway, the primary focus of guidelines and CLASS is on the application of respectful care principles to stillbirth management, emphasising shared decision-making as the core of the entire process. This approach ensures that every parent is treated with respect and that any cultural aspects are duly considered and respected.

In addition, our findings may not reflect the experiences of parents who did not have access to the internet or who were not reached by the call of CiaoLapo organisation.

Apart from the will of healthcare professionals, some issues could hamper the enhancement of international guidelines in the Italian context and this aspect will be the object of further research, focused on the perception of healthcare professionals about the implementation of international guidelines.

Finally, future research should include more diverse and representative samples of parents as well as multiple methods and sources of data collection, to provide a more comprehensive and accurate picture of bereavement care after stillbirth.

## 6 Conclusion

Our study offers significant insights into parents’ experiences of bereavement care following stillbirth. In Italy, adherence to international guidelines was found to be low and varied across regions and healthcare units. A higher adherence to these guidelines correlated with increased satisfaction and perception of respectful care, as well as reduced levels of grief and posttraumatic stress symptoms. Our research suggests that healthcare professionals should adhere to established guidelines on bereavement care. Additionally, utilising the CLASS checklist as an easy-to-use, standardised tool can help assess and improve the quality of care, as it has been validated for satisfaction and parents’ psychological well-being. This study highlights the necessity for official national guidelines on bereavement care after stillbirth in countries that currently lack them, such as Italy. Furthermore, adequate training and support for healthcare professionals involved in perinatal care should be prioritised.

## Data Availability

All data produced in the present study are available upon reasonable request to the authors

