## Supplementary figures and images for "CLASS - CiaoLApo Stillbirth Support checklist: adherence to stillbirth guidelines and women’s psychological well-being"

### Supplementary figure 1

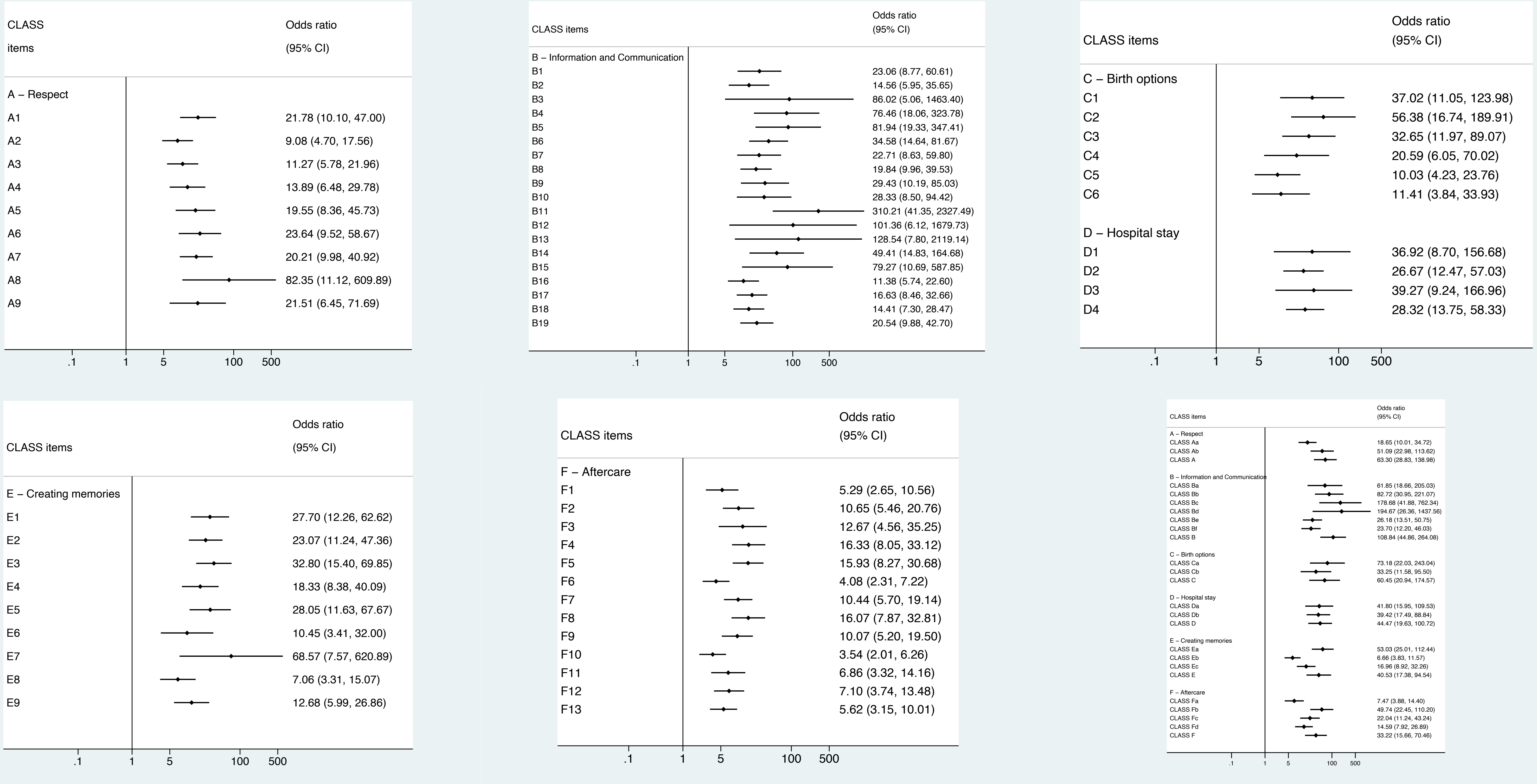
