## Supplementary table 1 for "CLASS - CiaoLApo Stillbirth Support checklist: adherence to stillbirth guidelines and women’s psychological well-being"

#### A - Respect

##### Aa - Baby

- ☐ A1 Name the baby
- ☐ A2 Touch, bath and dress the baby
- ☐ A3 Take pictures
- ☐ A4 Put the baby in a cot in the same room of the parents

##### Ab - Parents

- ☐ A5 Use a recognisable symbol for the staff, to denote bereavement
- ☐ A6 Acknowledge cultural, religious and spiritual needs of parents
- ☐ A7 Make repeated offers for holding the baby
- ☐ A8 Provide privacy (but not abandoning)
- ☐ A9 Allow women to spend time with their partner (or person of choice) before and after birth

#### B -Information and Communication

##### Ba - Timing

- ☐ B1 Discuss issues at most appropriate time
- ☐ B2 Choose a quiet and private environment
- ☐ B3 If the mother is unaccompanied, offer to contact the partner or a friend

##### Bb - Delivery of information

- ☐ B4 Be clear, honest, and sensitive
- ☐ B5 Repeat important information
- ☐ B6 Assure parents that it is normal to feel uncomfortable
- ☐ B7 Do not avoid questions

##### Bc - Mode of information

- ☐ B8 Give written information
- ☐ B9 Be comfortable showing emotions
- ☐ B10 Answer questions honestly
- ☐ B11 Listen to parents
- ☐ B12 Do not argue with parents

##### Bd - Terminology

- ☐ B13 Use parent-friendly language (simple and straightforward)
- ☐ B14 Use sensitive terms (avoid dehumanising terms as ‘fetus’, ‘product of conception’ etc)
- ☐ B15 Do not use medical jargon

##### Be - Burial and funeral services

- ☐ B16 Inform parents on laws and regulations
- ☐ B17 Provide parents help with funeral arrangements

##### Bf - Post-mortem examination

- ☐ B18 Give verbal and written information on autopsy/histology
- ☐ B19 Allow time for discussion issues regarding autopsy/histology

### C - Birth options

##### Ca -Timing

- ☐ C1 Inform in advance parents of factors relating to birth (when, how, etc)
- ☐ C2 Allow parents time to make appropriate decisions regarding birth

##### Cb - Mode of birth

- ☐ C3 Offer parents choices in birthing options (when available)
- ☐ C4 Offer parents the option of returning home (if possible)
- ☐ C5 Always offer obstetric analgesia
- ☐ C6 Avoid using sedation

### D - Hospital stay

##### Da - Environment

- ☐ D1 Offer parents the choice of the less distressing ward (possibly provide a private room)
- ☐ D2 Always allow to spend time with the baby

##### Db - After-birth care

- ☐ D3 Do not urge parents to leave the hospital
- ☐ D4 Offer continuity of care

### E - Creating memories

##### Ea - Spending time with baby

- ☐ E1 Clearly explain parents the length of time they are allowed to spend with their baby
- ☐ E2 Let the parents all the time they need to stay with the baby
- ☐ E3 Always offer parents to see and hold the baby, but do not force them; discuss what is the best option for them
- ☐ E4 Respect their wishes regarding seeing and holding the baby
- ☐ E5 Prepare parents for the appearance of the baby

##### Eb - Parenting

- ☐ E6 Allow parents to bath and dress their baby
- ☐ E7 Allow siblings to spend time with the baby

##### Ec - Mementos

- ☐ E8 Take pictures of the baby and keep them in the hospital
- ☐ E9 Provide parents with mementos (lock of hair, hand and footprints, ID bracelet, cot card, pictures)

### F - Aftercare

##### Fa - Spiritual / religious needs

- ☐ F1 Allow parents to bless, baptize or perform any other religious ceremony for their baby

##### Fb - Maternal changes

- ☐ F2 Inform mothers on milk production
- ☐ F3 Offer a consultation with a lactation consultant

- ☐ F4 Inform mothers on bleeding
- ☐ F5 Inform mothers on psychological consequences of perinatal loss
- ☐ F6 Provide a follow-up at 6 weeks post birth. Give name and number of the case manager

#### Fc - Support services

- ☐ F7 Provide early psychological support
- ☐ F8 Provide written information on support services (include fathers)

#### Fd -Follow-up

- ☐ F9 Provide name and number of the case manager
- ☐ F10 Provide an appointment with a senior obstetrician within 2 months
- ☐ F11 Where possible, use rooms away from the hospital
- ☐ F12 Give and explain results of post-mortem examinations
- ☐ F13 Discuss implication for future pregnancies
