## Supplementary table 2 for "CLASS - CiaoLApo Stillbirth Support checklist: adherence to stillbirth guidelines and women’s psychological well-being"

|  | **No.** | **%** |
| --- | --- | --- |
| **Age** |  |  |
| < 35 y | 94 | 36.0% |
| 35 - 40 y | 91 | 34.9% |
| > 40 y | 76 | 29.1% |
| **Level of education** |  |  |
| Lower secondary | 12 | 4.6% |
| Upper secondary | 118 | 45.2% |
| First stage tertiary | 104 | 39.8% |
| Second stage tertiary | 27 | 10.3% |
| **Voluntary ToP** |  |  |
| No | 222 | 86.0% |
| Yes | 36 | 14.0% |
| **Months after loss** |  |  |
| <12 months | 66 | 25.3% |
| 12-30 months | 65 | 24.9% |
| 30-60 months | 65 | 24.9% |
| >60 months | 65 | 24.9% |
| **Gestational age at loss** |  |  |
| 20 - 25 wks | 87 | 33.3% |
| 26 - 36 wks | 92 | 35.2% |
| 37 - 42 wks | 82 | 31.4% |
| **Subsequent pregnancies** |  |  |
| No | 120 | 46.0% |
| Yes | 141 | 54.0% |
| **Currently pregnant** |  |  |
| No | 233 | 89.3% |
| Yes | 28 | 10.7% |
| Total | 261 | 100.0% |

|  | **No.** | **%** |
| --- | --- | --- |
| **Medical history** |  |  |
| Diabetes | 12 | 4.6% |
| Coeliac disease | 7 | 2.7% |
| Obesity | 38 | 14.6% |
| Autoimmune disease | 21 | 8.0% |
| Coagulopathy | 30 | 11.5% |
| **Psychological history** |  |  |
| Anxiety | 61 | 23.4% |
| Depression | 22 | 8.4% |
| Bipolar Disorder | 1 | 0.4% |
| Eating Disorders | 10 | 3.8% |
| **Number of losses** |  |  |
| 1 loss | 190 | 72.8% |
| 2-3 losses | 45 | 17.2% |
| >3 losses | 26 | 10.0% |
| Total | 261 | 100.0% |
